# Differences in Treatment and Outcome of Patients with ST- Elevation Myocardial Infarction (STEMI) and Non-STEMI in Germany

**DOI:** 10.64898/2026.02.13.26346292

**Authors:** SA Lange, C Engelbertz, L Makowski, P Dröge, T Ruhnke, C Günster, J Gerß, H Reinecke, J Koeppe

**Author notes:** <u>Address for correspondence as well as requests for reprints:</u>* PD Dr. med. Stefan A. Lange, Dept. of Cardiology I - Coronary and Peripheral Vascular Disease, Heart Failure, University Hospital Muenster, Albert-Schweitzer-Campus 1, D-48149 Muenster, Germany.

## Abstract

**Background:** Although ST-segment elevation myocardial infarction (STEMI) and non-STEMI (NSTEMI) are very similar regarding pathophysiology and clinical treatments, especially NSTEMI comprises a much more heterogenic group of patients and underlying diseases. We therefore aimed to assess the treatments and outcomes of both entities in a large contemporary cohort.

**Methods:** Patients with STEMI and NSTEMI between 01/2010 to 12/2018 were identified from the largest German Health Insurance (AOK, ≈26 million members). Patient demographics, their hospital course, adherence to guideline-directed drug therapy and overall survival were assessed.

**Results:** In total 544,529 patients (mean age 74, IQR 62-82), one third of whom had a STEMI. Chronic kidney disease, peripheral arterial disease, and heart failure were more common in patients with NSTEMI. Patients with STEMI were more likely to get coronary angiograms and percutaneous coronary interventions. Although STEMI more frequently led to cardiogenic shock, the rate of serious cardiac events was lower. Mortality was higher for STEMI only within the first 30 days, whereas long-term survival rates were better. The combination of statins, angiotensin converting enzyme inhibitors /angiotensin receptor blockers, beta blockers, and oral anticoagulants or antiplatelet agents was associated with higher overall survival in patients with STEMI (hazard ratio [HR] 0.20; 95% confidence interval [95%CI] 0.18 – 0.24; p<0.001) or NSTEMI (HR 0.30; 95%CI 0.28 – 0.33; p<0.001). Nevertheless, the prescription rates decreased over time, particular in patients with NSTEMI.

**Conclusion:** Clear differences between STEMI and NSTEMI were observed regarding short-and long-term survival. Guideline-recommended therapy improved long-term survival, but decreased during the follow-up period.

## Introduction

The guidelines of the European and American societies of cardiology for the treatment of acute coronary syndrome (ACS) make a clear distinction between ST-segment elevation-myocardial infarction (STEMI) and non-STEMI (NSTEMI) (**1, 2**). In terms of cause and treatment, STEMI and NSTEMI are considered separately. STEMI generally receives more attention and a delay in targeted treatment, in most cases by percutaneous coronary intervention, leads to a dramatic increase in mortality. If untreated, the 30-day mortality rate for STEMI is between 15.7% and 24.6% (**3**). The situation appears to be different for the more common NSTEMI. Patients with these infarcts receive coronary angiography less frequently and PCI is also used less often (**4**). Lysis therapy is not indicated. There are also differences in drug therapy between STEMI and NSTEMI. While patients with STEMI are often prescribed dual antiplatelet therapy for a period of 12 months after percutaneous coronary intervention, shorter treatment intervals are often considered justified for patients with NSTEMI (**5**). Guideline-recommended drug therapy with cholesterol synthesis enzyme (CSE) inhibitors and angiotensin-converting enzyme inhibitors (ACEI) or angiotensin receptor blockers (ARBs) among others, reduces mortality and lowers the major adverse cardiac event (MACE) rate, especially after a STEMI (**6**). The extent to which there are also relevant differences in the frequency of prescription to medication in patients with NSTEMI or STEMI is of great interest.

Therefore, we aimed to assess the medical treatment with guideline-recommended medication and the outcome in patients with NSTEMI and STEMI using the longitudinal data of a large German health insurance of the years 2010 – 2019.

## Methods

Reimbursement of services in the German healthcare system is based on diagnoses and the severity of the illness. In order to determine this, it is necessary to define the respective diagnoses individually using the “German Diagnosis Related Groups” (G-DRG) diagnosis key. In doing so, a main diagnosis and an unlimited number of secondary diagnoses are determined for the respective patient during their inpatient stay. This procedure allows the determination of comorbidities and complications. The basis for the coding is the “International Statistical Classification of Diseases and Related Health Problems, 10th Revision, German Modification” (ICD-10 GM). In addition, all diagnostic, interventional and surgical procedures are coded according to the German Procedure Classification (OPS).

The patient selection, data source and data accessibility used for this study, and the ethical evaluation, can be found in the supplements. The ICD-10 GM, OPS and Anatomical Therapeutic Chemical/Defined Daily Dose Classification-ATC codes used are summarized in supplemental **Table S1**.

As data source we used health claims data from eleven legally independent statutory health insurance funds of the “AOK – Die Gesundheitskasse” (local health care funds). Enrolment to statutory health insurance is unrestricted regarding age, health status, income, employment, or geographical region. In brief, all patients aged ≥18 years with NSTEMI and STEMI-ACS as their main diagnosis, who were insured by the AOK and admitted to hospital during the index period from January 1, 2010 to December 31, 2018, were included in the database analysis. The primary objective of the study was to determine differences in patient characteristics, treatment strategies and outcomes between NSTEMI-ACS and STEMI-ACS patients.

### Outcome measures

The primary endpoints included short survival and long-term survival, with long-term survival only including patients who had survived for 90 days.

### Statistics

Multivariable Cox regression with time-dependent covariables were used to evaluate differences between STEMI vs NSTEMI for all endpoints. Moreover, to account for different treatment effects for guideline-based drug therapy on survival between STEMI and NSTEMI patients, an interaction term between medication and type of infarction was added. The statistical methods in detail can be found in the supplemental material (statistical methods). Moreover, charges of the first hospitalization were analyzed descriptively.

## Results

The total cohort of patients with an acute myocardial infarction, STEMI and NSTEMI, in this 9-years period comprised 544,529 patients, with mean age 74 (interquartile range [IQR] 62 – 82 years). One third of these patients were admitted to hospital with STEMI (181,741 or 33.4%). The median age of patients with STEMI was 8 years younger than patients with NSTEMI (68, IQR 57 – 78 vs. 76, IQR 65 – 83 yrs.). The proportion of women among AMI patients with acute myocardial infarction (AMI) was significantly lower than that of men (39.2%). The difference was particularly pronounced in the STEMI cohort (STEMI 35.0% vs. NSTEMI 41.34%, p <0.001).

### In-hospital treatment

The complete baseline data and in-hospital outcome results are presented in **Table 1A and 1B**. Baseline characteristics were different between patients with STEMI or NSTEMI (see **Table 1A**).

**Table 1:**
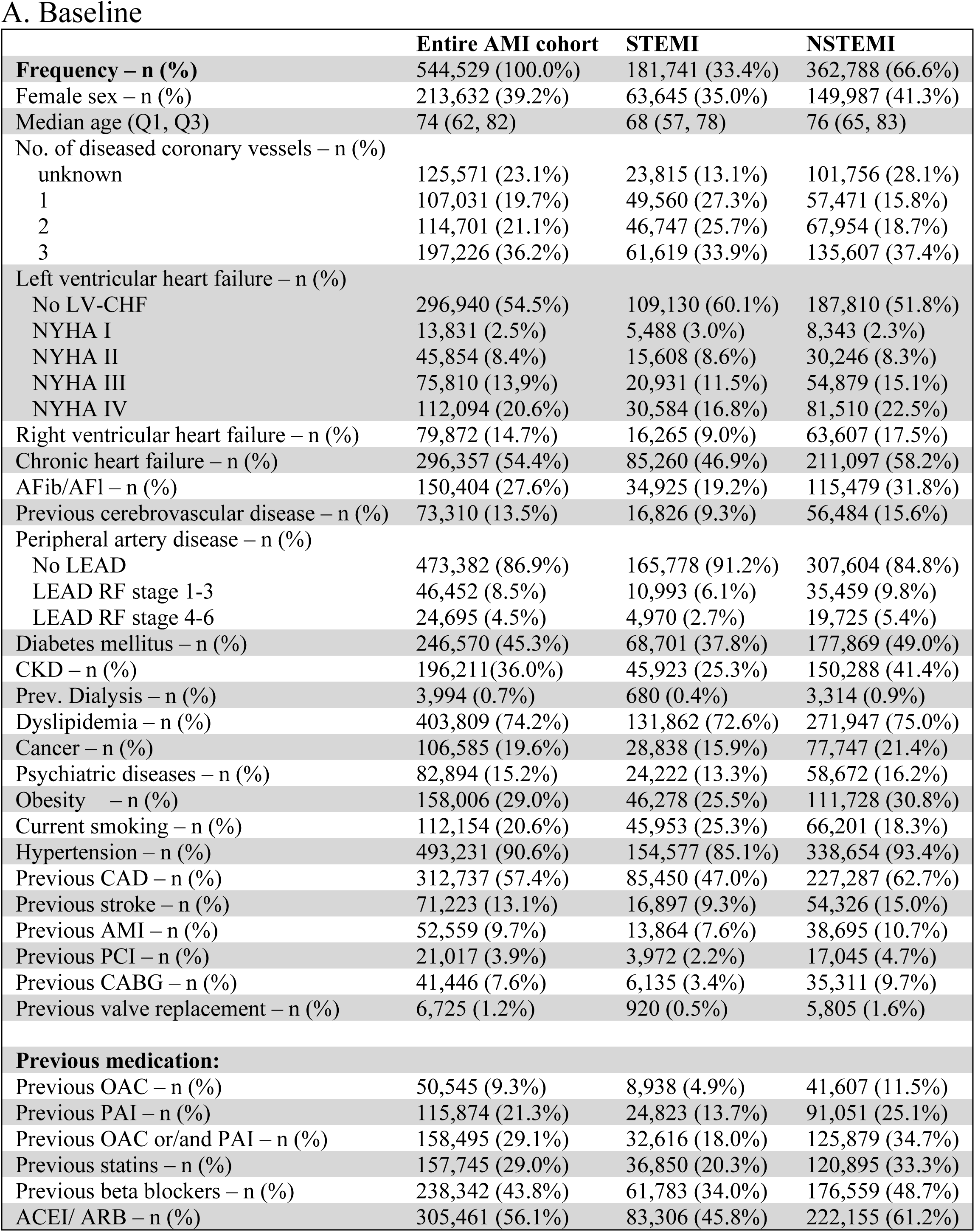
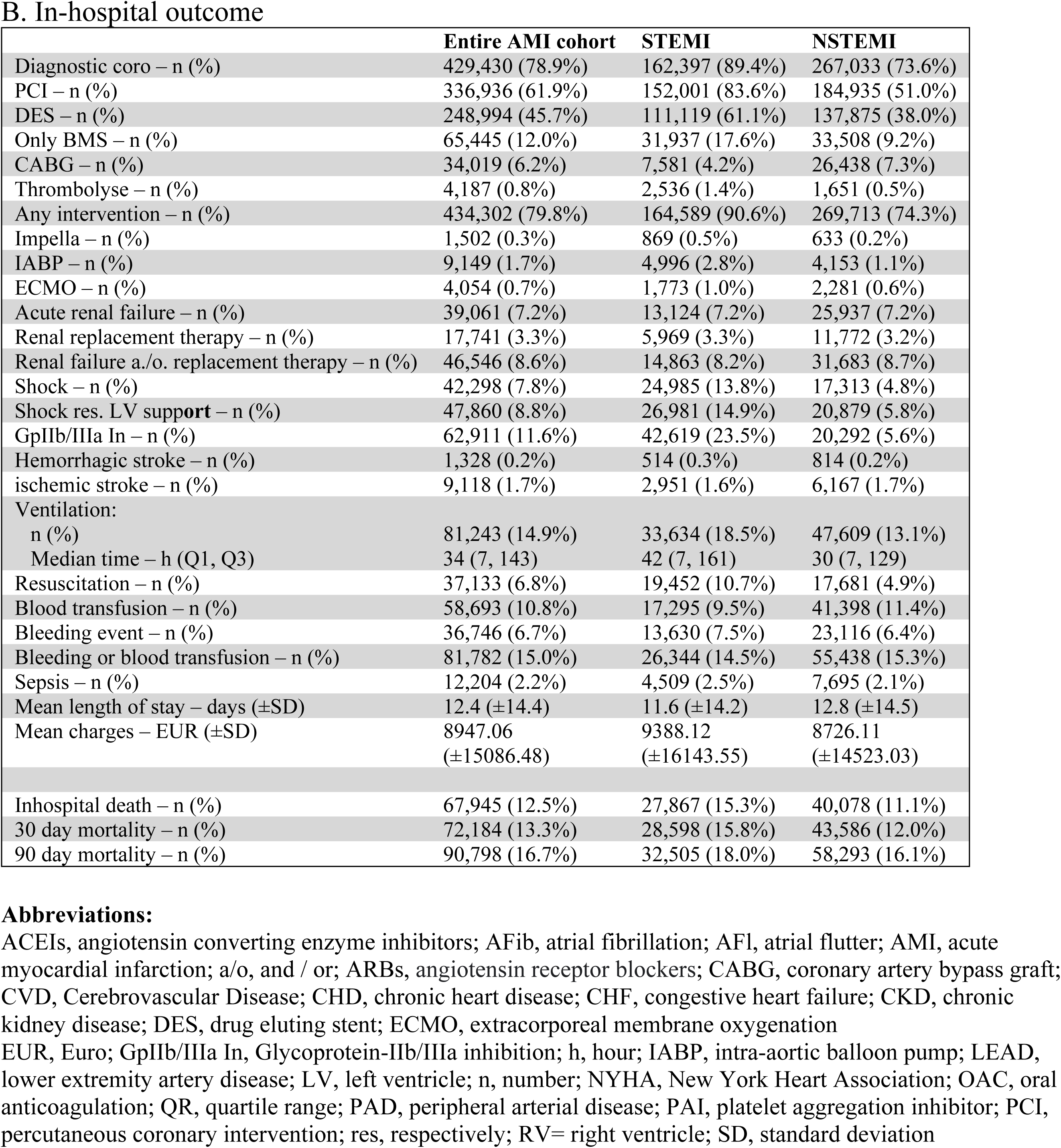
Baseline characteristics for STEMI vs. NSTEMI.

With regard to the diagnostics and therapies performed, patients with STEMI were more likely to receive diagnostic coronary angiography (STEMI 89.4% vs. NSTEMI 73.6%, p <0.001) and percutaneous coronary interventions (STEMI 83.6% vs. NSTEMI 51.0%, p <0.001) and less likely bypass surgery than patients with NSTEMI (STEMI 4.2% vs. NSTEMI 7.3%, p <0.001; see **Table 1B**). Patients with STEMI were more likely to be in a life-threatening situation (**Table 1B**). In detail: 19,452 patients with STEMI (10.7%) vs. 17,681 (4.9%) patients with NSTEMI required resuscitation. 26,981 (14.9%) vs. 20,879 (5.8%) had a shock and/ or required left ventricle (LV) live support. Of these 37,133 patients with shock (STEMI or NSTEMI), 14,705 (rep. 39.6%) patients received mechanical circulation support. Of these, 1,502 patients received an axial pump (10.2%) (Impella®), 9,149 patients received intra-aortic balloon pumps (IABP, 62.2%) and 4,054 received extracorporeal membrane oxygenation (ECMO, 27.6%). In line with the distribution of cardiogenic shock, patients with STEMI were more likely to require ventilation support (18.5% vs. 13.1%, p < 0.001) and also had a longer median ventilation time (42 h, IQR 7–161 h vs. 30 h, IQR 7–129 h). Further in-hospital complications are presented in **Table 1B**. Moreover, the costs for STEMI treatment during the index hospitalization were in mean about €650 higher than the treatment costs for NSTEMI (€9,388 ± 16,144 vs. €8,726 ± 14.523).

### Overall survival

While a higher mortality rate was observed in patients with STEMI in the acute phase (30-day mortality: STEMI 15.8% vs 12.0% in NSTEMI patients), the long-term course changes (see Figure 1). Three-years after AMI, 28.2% (95% confidence interval [95%CI] 28.0 – 28.4%) of patients with STEMI but 37.0% (95%CI 36.9 – 37.2%) of patients with NSTEMI were deceased. Unadjusted event rates for the endpoints reinfarction or death and major adverse cardiac and cerebrovascular events (MACCE) are presented in **Table S2**. This effect was confirmed by a multivariable landmark analysis (see **Table S3**). Within the first month after AMI, STEMI was associated with higher risk of death compared to NSTEMI (hazard ratio [HR] 1.84; 95%CI 1.81 – 1.87; p<0.001; see **Table S4**), while the opposite effect was observed later in course, i.e. after one month, STEMI was associated with lower risk of death (HR 0.81; 95%CI 0.80 – 0.82; p<0.001) compared to NSTEMI. As a sensitivity analysis, the association of infarction type (STEMI or NSTEMI) and survival was further analyzed using multivariable Cox regression with time-dependent variables, showing similar effects as observed in the landmark analysis (see **Figure S1**).

**Figure 1.**
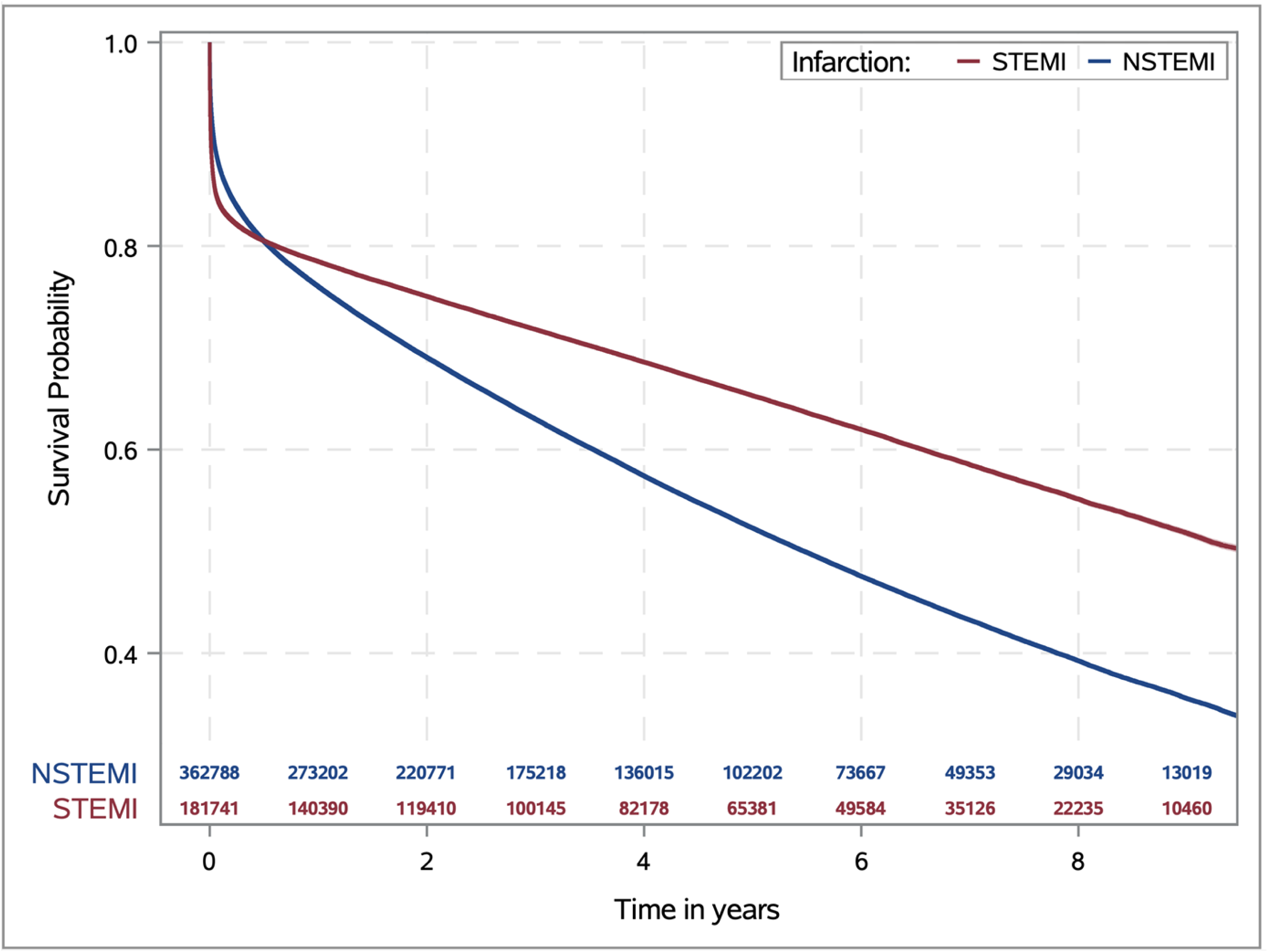
Survival probability comparing STEMI vs NSTEMI over time after infarction. Within the first six months after the first AMI, the survival advantage of patients with NSTEMI over patients with STEMI was already reversed. **Abbreviations:** NSTEMI, non-ST-segment elevation myocardial infarction; STEMI, ST-segment elevation myocardial infarction.

### Effect of guideline-directed drug therapy on mortality

A decisive factor that has a direct influence on the survival of patients with AMI is adherence to the prescription of the four typical medications, i.e., oral anticoagulants (OACs) or antiplatelet agents (PAIs), statins, beta blockers, angiotensin-converting enzyme inhibitors (ACE-I) or angiotensin receptor blockers (ARBs), following myocardial infarction. The multivariable Cox regression analysis for the use of OAC/PAI, statins, betablockers and ACEI/ARB to reduce mortality after STEMI or NSTEMI showed a clear benefit for each of the 4 drug groups (all p <0.05; see **Figure 2 upper panel**). Furthermore, risk reduction increased with the number of specific drug groups in patients with NSTEMI or STEMI, while the association was more pronounced in patients with STEMI (p <0.05, see **Figure 2 lower panel**). In detail, a guideline-recommended prescription using all four drugs was associated with a lower risk of death in patients with STEMI (HR 0.20; 95%CI 0.18 – 0.24; p<0.001) and in patients with NSTEMI (HR 0.30; 95%CI 0.28 – 0.33; p<0.001; see **Figure 2**).

**Figure 2:**
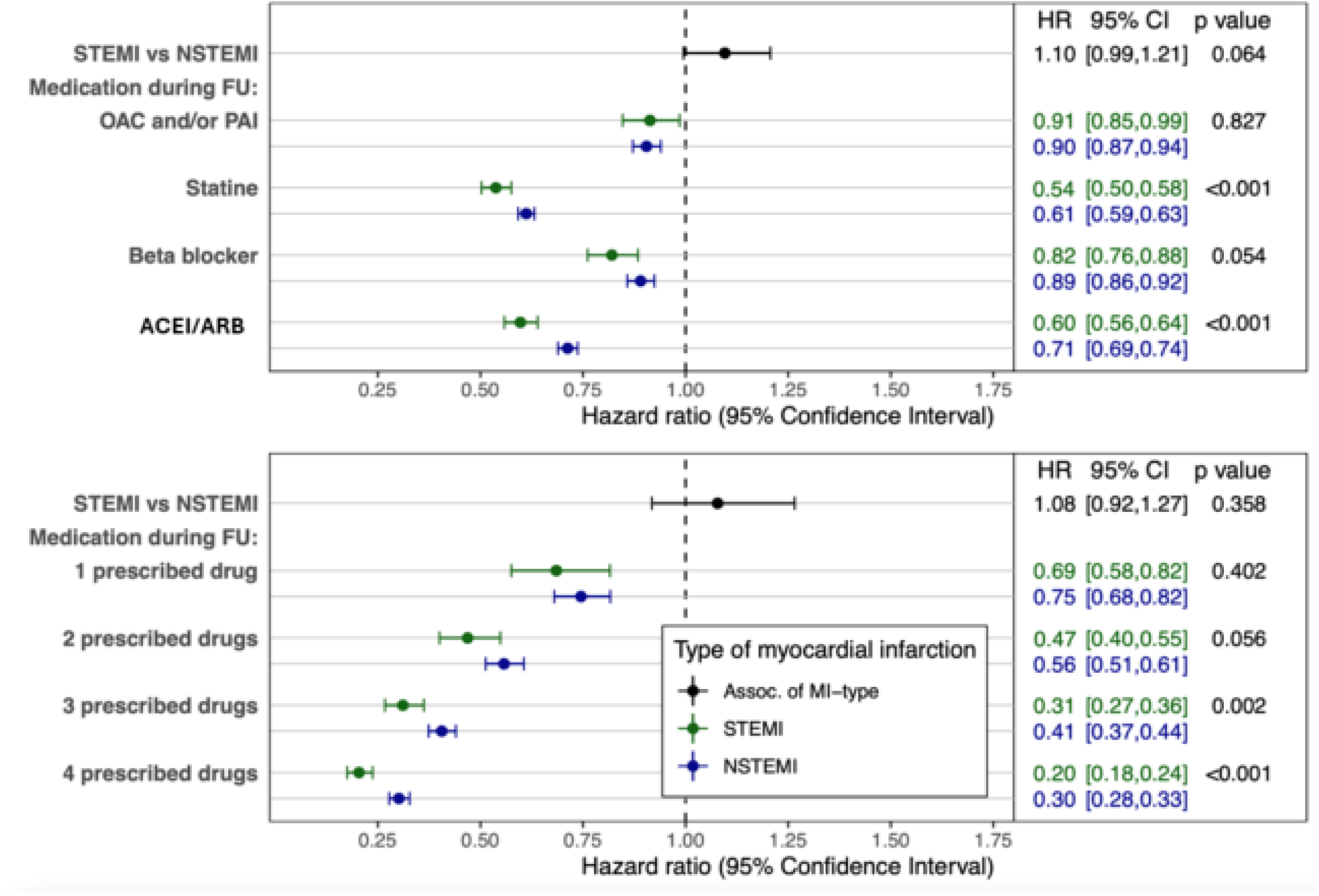
Cox Regression for overall survival including guideline-based medication in interaction with the type of infarction (upper panel) and including only the number of prescribed drugs (lower panel). **Abbreviations:** AMI, acute myocardial infarction; ACEI/ARB, angiotensin-converting-enzyme inhibitors / angiotensin II receptor blocker; CI, confidence interval; FU, follow up; HR, hazard ratio; NSTEMI, non-ST-segment elevation myocardial infarction; STEMI, ST-segment elevation myocardial infarction.

Prescription rates for the four cardiovascular medications recommended in the guideline declined in the years following an AMI, even though adherence to the prescription has been shown to improve survival rates (see **Figure 3** and **Table S4**). For example, only 42.3% (95% CI 42.0–42.6%) of patients with STEMI and 36.8% (95% CI 36.6–37.0%) of patients with NSTEMI who were still alive three years after an AMI received guideline-recommended drug therapy with all four drugs (see **Figure 3**).

**Figure 3.**
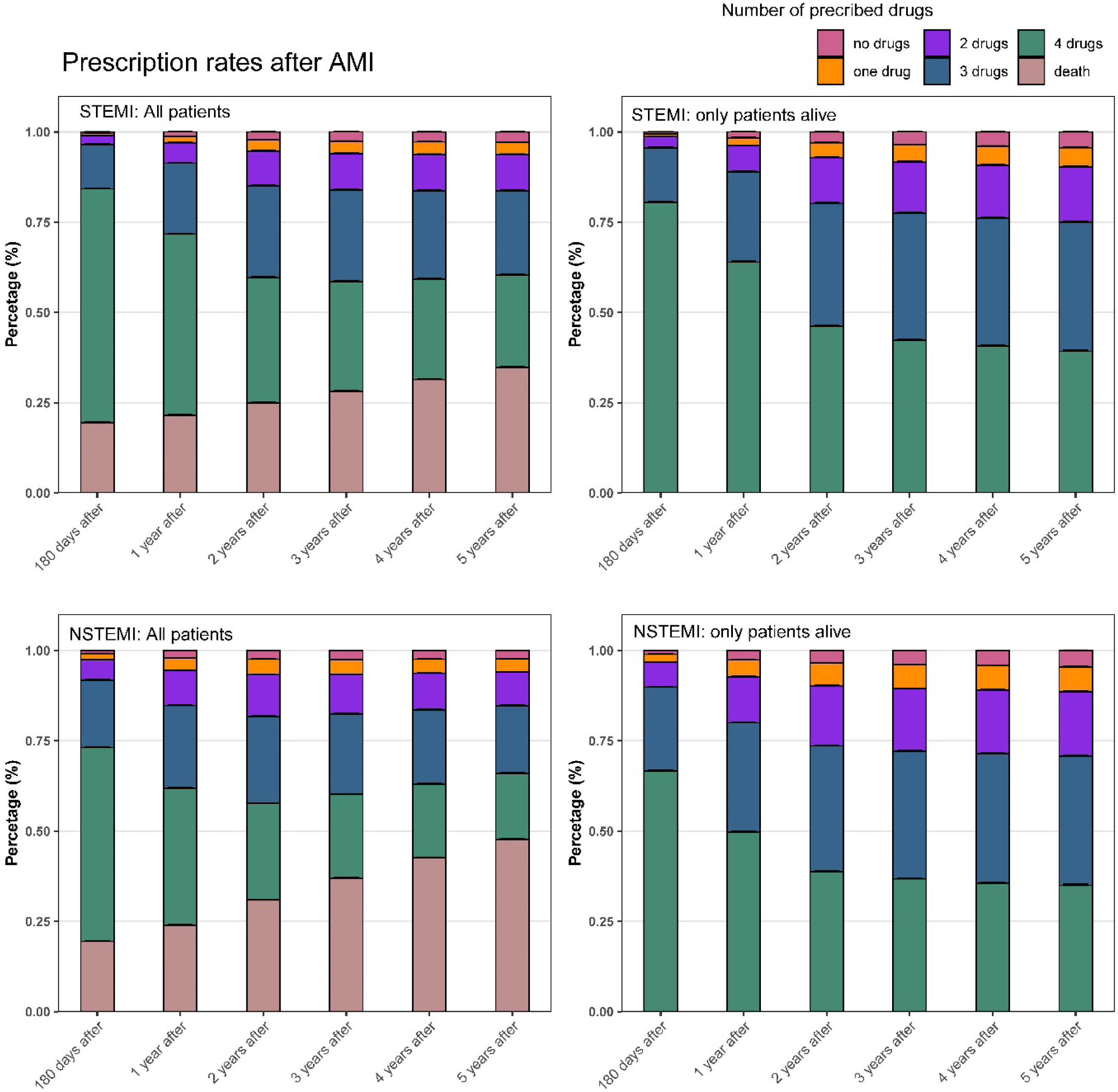
Declining prescription rates for the most important drugs after AMI Prescription rates up to five years after AMI. Upper panel shows prescription rate in STEMI patients, on the left side all patients and on the right side only AMI survivor, lower panel shows prescription rate in NSTEMI patients, on the left side all patients and on the right side only AMI survivor. Prescription rates were given by the actual state probability determined using multi-state model. **Abbreviations:** AMI, acute myocardial infarction; NSTEMI, non-ST-elevation myocardial infarction; STEMI, ST-elevation myocardial infarction.

## Discussion

The presented study included patient data from more than 26 million insured persons of the general local health insurance funds (AOK) in Germany over a period of 9 years. Of these insured persons, approximately 550,000 patients with myocardial infarction, STEMI and NSTEMI, were hospitalized between January 1, 2010 and December 31, 2018 and subsequently followed up for up to nine years for survival. Serious differences between patients with STEMI or NSTEMI were documented regarding outcome and medical treatment. The higher mortality in patients with STEMI in the early phase after the event should be emphasized, whereas this disadvantage shifted to the disadvantage of patients with NSTEMI after one month and then remained constant at this level over the entire follow-up period. Compared to the European countries Sweden, Norway, Estonia and Hungary, the 30-day mortality rate for patients with STEMI in Germany was higher (15.3%) than for patients with STEMI in Sweden (9.9%), Norway (12.4%), Estonia (13.4%), and Hungary (13.3%) (**7**). The 30-day mortality rate for patients in Germany with NSTEMI was 12.0% in our data set. This is only slightly higher than the comparative values in Hungary (11.8%), Norway (10.9%), and Estonia (10.2%). However, it was higher than the 1-month mortality data for NSTEMI in Sweden (5.2%) (**8**). It should be noted that patients over 80 years of age were underrepresented in the corresponding SWEDHEART registry (**8**). Older data collected between 2003 and 2013 in the United Kingdom (UK) for AMI showed a 1-month mortality rate of 8% for STEMI and 6.7% for NSTEMI in the UK (**9**). In France, the 30-day mortality rate in 2015 was around 3% for both STEMI and NSTEMI (**10**).

However, it is generally difficult to compare the statistics collected from different regions and then use them to draw conclusions about the quality of treatment. Cardiovascular risks, the type of AMI treatment and the resulting post-hospitalization survival rates for patients with STEMI or NSTEMI vary greatly from region to region (**11,12**).

The comparability of mortality results must also be critically examined because there are differences between individual countries in the way patient data is recorded. In particular, the recording of so-called ‘day cases’ is not uniformly regulated (**13**). Our data consists of secondary analyzed individual patient data and not patient cases. Multiple recording of the same person or non-recording of stays of less than one day (i.e. short stays or outpatient cases) is excluded. Nevertheless, it is necessary to address the possible reasons for the difference in survival between STEMI and NSTEMI. Along with the increased mortality in patients with STEMI, shock events occurred more frequently in the early phase of STEMI. In contrast, patients with NSTEMI were more likely to have signs of chronic heart failure and had more findings of coronary 3-vessel disease. In addition, serious comorbidities relevant to long-term survival were documented more frequently in patients with NSTEMI, e.g., advanced renal insufficiency, cerebrovascular disease, atrial fibrillation and PAD. The observation that patients with STEMI had different short- and long-term survival rates compared to patients with NSTEMI has already been described in several studies and does not appear to have changed significantly in the past two decades. A considerable number of studies from various high-income countries have shown improved long-term survival for patients with STEMI (**14–19**). However, it could be shown when relevant comorbidities are taken into account, the differences between STEMI and NSTEMI patients appear to disappear in terms of long-term survival, while patients with STEMI remain disadvantaged in terms of short-term survival (**20, 21**).

Patients with NSTEMI underwent fewer invasive coronary diagnostics procedures and consequently received fewer PCI procedures. However, it should be mentioned at this point that coronary intervention was not necessarily associated with a survival advantage, particularly in older NSTEMI patients (**22**). Another reason for the poorer long-term survival of patients with NSTEMI may also lie in the interventional treatment strategy for these patients between 2010 and 2018. As a result, complete revascularization of the coronary vessels was often not attempted, especially in older patients with comorbidities such as renal failure and a high risk of bleeding. Before the studies demonstrating the advantages of complete revascularization, the motto “culprit lesion only” was often preferred, particular in patients with NSTEMI. However, it has recently been shown that early complete revascularization generally offers a survival advantage for patients with AMI (**23, 24**). It should be noted at this point that it has recently been shown that CABG surgery, which is known to aim for complete revascularization, has a lower long-term mortality rate in patients with NSTEMI compared to PCI. However, patients with life-limiting comorbidities such as cancer, terminal renal insufficiency, stroke or dementia were excluded from the analysis (**25**). Nevertheless, acute bypass surgery generally plays a minor role in STEMI or NSTEMI. Here too, no relevant difference could be documented between the probability of bypass surgery in STEMI (4.2%) and NSTEMI (7.3%) compared to the other countries mentioned above.

Beyond the initial hospital stay, there are other significant factors that influence the survival rate after AMI. In a direct comparison of patients with AMI in Germany, the United Kingdom, the Netherlands, Sweden, and Norway, patients in Germany suffer more frequently from high blood pressure, diabetes mellitus, and higher LDL cholesterol levels. (**7, 8, 26**). Accordingly, a key focus of the study presented here was also on the pharmacological follow-up treatment of myocardial infarction. It was thus demonstrated that the use of the four classic drugs for treating CCS (ACEI/ARB, beta blockers, statins and PAI/OAC) has synergistic effects on reducing mortality. Taken together, they reduce the risk of death after a STEMI by approx. 80 % and after a NSTEMI by approx. 70 %. Nevertheless, the frequency of prescriptions and adherence to medication in patients with STEMI or NSTEMI decreased over time (**6, 27**). This is particularly emphasized in the use of lipid-lowering cholesterol synthesis enzyme (CSE) inhibitors. For example, the prescription frequency of statins fell by 13 percentage points from just under 93% initially in STEMI patients to just under 80% after 5 years. In patients with NSTEMI, the already initially lower prescription rate of CSE inhibitors fell from just under 83% to just under 73% after 5 years. Unfortunately, our study did not allow us to ask about the individual reasons for this. However, it is not only the prescription of CSE inhibitors themselves that plays a role in lowering LDL cholesterol, but also the dose of statins and, if necessary, supplementary lipid-lowering drugs. The LDL cholesterol value is primarily used as a guideline. Comparisons of treatment practices for patients with stable coronary heart disease between different countries in the DYSIS II ACS study from 2017 showed that ACS patients in Germany still had a high prescription rate of statins 120 days after the primary ACS event (91.5%) (**28**). However, LDL laboratory tests were only performed in 27.9% of cases. This may be one reason why the LDL target value (at that time <70 mg/dl) in Germany was only achieved in 19.3% of patients. By comparison, in France, the lipid profile was determined in > 50% of cases during the course of treatment, and the target values were achieved by 50.6% of patients (**28**). One reason for this may be the differing recommendations issued by professional associations in Germany. While the German Society of Cardiology (DGK) aims for LDL target value-oriented secondary prophylaxis in line with the ESC guideline, the national care guideline (Nationale-Versorgungs-Leitlinie, NVL) for CCS, which is usually followed by general practitioners providing follow-up treatment, does not recommend this. Instead, it advises taking a fixed dose of a CSE inhibitor as a primary treatment, regardless of the actual reduction in LDL (**29, 30**).

In summary, it can be said that in order to further improve survival rates after MI in Germany, it is necessary to consistently implement the integration of inpatient interventional/surgical therapy on the one hand and outpatient risk-adjusted secondary prophylaxis with medication in accordance with the guidelines for the treatment of ACS on the other. It is essential to inform patients about their individual risks and treatment options.

### Strength and limitations

The analyzed data base from the AOK covers around one third of the German population. However, since no data from other insurance companies has been received, selection bias cannot be entirely ruled out.

Even if there remains a residual risk of coding errors, it can be assumed that the validity of the entry of ICD and OPS codes is likely to be high, as the MD (*Medizinischer Dienst*/Medical Service) of the health insurance funds checks up to 30% of cases for accuracy and the coding has a direct influence on hospital reimbursement.

The analysis is subject to general limitations, which are also related to a retrospective study design based on health claims data which were originally generated and collected for billing purposes. In addition, information on the clinical condition of individual patients, treatment procedures (e.g. door to balloon time), echocardiographic findings and laboratory parameters (e.g. maximum creatine kinase, troponin (T/I) and nt-Pro-BNP) is missing, making a more in-depth analysis of individual cases impossible. In addition, patients who died outside the hospital before admission due to a heart attack could not be included in the data analysis.

Medication adherence was determined using pharmacy records and not by patient interview or direct measurement of drug or metabolite concentrations in blood or urine. We did not have clinical information on the severity of cardiovascular disease, lifestyle factors and specific contraindications or potential adverse drug reactions that may influence the prescription and early discontinuation of medication. Finally, we cannot rule out with certainty that changes in the Universal Diagnostic of Myocardial Infarction (UDMI) and changes in the AHA and ESC guidelines for the diagnosis and treatment of myocardial infarction during the observation period could have an impact on our results.

## Conclusion

In patients with ACS, there are clear differences between STEMI or NSTEMI in terms of short-and long-term survival. Guideline-based therapy improved long-term survival but adherence declined in patients with STEMI or NSTEMI during follow-up.

## Contributors

The authors thank the personnel of the AOK research institute for their technical support.

## Data Availability

Due to the Federal Data Protection Act (BDSG), the data used in this study cannot be made available in the manuscript, in the supplementary files or in a public repository. Therefore, they are stored on a secure drive at the AOK Research Institute (WIdO) to facilitate replication of the results. In general, access to statutory health insurance data for research purposes is only possible under the conditions laid down in the German Social Code (SGB V § 287). Requests for data access can be submitted as a formal application to the relevant data protection authority, specifying the recipient and the purpose of the data transfer. Access to the data used in this study can only be granted to external parties under the conditions of the cooperation agreement of this research project and after written approval by the health insurance company. For assistance in obtaining access to the data, please contact.

## Contributions statement

SA Lange and J Koeppe were involved to this work in conception of design, for intellectual content, analysis and interpretation of the data, drafting of the paper, and final approval of this version.

J Koeppe and J Gerß were involved in conception of design, analysis and interpretation of data, revising it critically and approval of the manuscript.

L Makowski and C Engelbertz were involved to this work in conception of design, for intellectual content, analysis and interpretation of data, and approval of the manuscript. J Koeppe, H Reinecke and C Engelbertz critically revised of the manuscript.

P Dröge, T Ruhnke, and C Günster were involved in provision of data, revising it critically for intellectual content; revising it critically and approval of the manuscript.

H Reinecke was involved to this work in conception of design, for intellectual content, analysis and interpretation of the data, final approval of this version, and financial support.

